# Musculoskeletal Advertising Focuses on Whites and Overlooks Minority Communities

**DOI:** 10.1101/2022.12.14.22283466

**Authors:** Kelsey A. Rankin, Robert John Oris, Adithi Wijesekera, Daniel H. Wiznia

## Abstract

**Introduction:** Demographic disparities in musculoskeletal (MSK) health exist in the US. Racial representation in advertising has been shown to influence buying patterns. By focusing advertising toward majority groups, direct-to-consumer advertising may exacerbate MSK disparities by neglecting underrepresented minorities. To better understand how race is represented across MSK advertisements and how this may influence patterns in MSK health, we reviewed advertisements in popular magazines, using online databases for collection.

**Methods:** 8 magazine types were chosen. Racial distribution was analyzed using Pearson’s chi-squared and chi-squared goodness of fit tests. Fisher’s exact test was used when >20% of cells had n<5. Significance was set at p<0.05.

**Results:** Of advertisements that featured a model, white models were overrepresented (p<0.001), and Hispanic and Asian models were underrepresented (p<0.001). Only 7.3% of advertisements featured multiple models of different races or ethnicities, while 92.7% did not. African American models were overrepresented as athletes (p<0.001) and underrepresented in pain relief ads (p<0.001).

**Discussion:** There is poor representation of minorities in MSK advertisements. Even when controlling for US population demographics, white models were overrepresented, and models of minority races are underrepresented. African American models were typecast as athletes and underrepresented in pain relief ads.

## Introduction

Racial and ethnic disparities in musculoskeletal (MSK) health exist in the U.S. The prevalence of hip and knee osteoarthritis is highest among African Americans and Hispanics [1–5]. African Americans have the highest burden of hip and knee osteoarthritis [2–4], as well as disproportionately high losses of quality-adjusted life years (QALYs), due to osteoarthritis [6].

Racial and ethnic differences in food insecurity [8], access to health-promoting foods [9], and engagement in exercise and healthy eating [10] exist in the U.S., and are likely contributory factors to these observed differences in MSK health.

Direct-to-consumer advertising represents an important means of exposure to health promoting products. It has been shown that racial minorities, including African Americans and Hispanics, have been historically underrepresented in advertising [11, 12]. Representation of African American models in magazine advertisements is increasing, but is skewed toward depicting African American models as athletes or musicians [11].

In addition, it has been shown that the race of models used in advertisements influences buying patterns: individuals are more likely to make a purchasing decision if they identify with the race and/or ethnicity of the model [13–19]. Magazines aimed at African American and Hispanic populations have been shown to display significantly fewer health-promoting advertisements and more health diminishing advertisements [20].

Examining racial diversity in MSK magazine advertising has important implications for product utilization and health disparities. The extent to which advertisements that promote musculoskeletal health are aimed at minority populations is unknown. This study aims to examine whether racial and ethnic disparities exist in MSK magazine advertising, to support a better understanding of how more diverse advertisement content may improve MSK disparities.

## Methods

### Study design

Advertisements relating to MSK health were collected from magazines, both digital and print. The following definition was developed to identify an MSK health advertisement: any advertisement whose product or service would benefit an individual’s orthopedic health (skeletal, cardiac, muscular, or nutritional). The following databases were used to obtain magazine content: RBdigital, ZINIO magazine database, *Magzter*, and Amazon.

Data was obtained from the eight following magazine types: women’s health, men’s health, food/dietary health, elderly health, sports health, general lifestyle, African American health, and Hispanic health. Each magazine type was populated with three magazines. The magazines chosen for each type were based on the highest number of subscriptions and accessibility to one complete year’s worth of issues from the above databases. A June 2019 industry whitepaper from the Alliance of Audited Media was utilized to determine magazines with the greatest subscription counts [23]. A complete list of magazine titles for each magazine type is included in Supplemental Table 1.

One year’s worth of magazines was examined, with the primary time frame being January-December 2019. However, within the four magazine sources that were utilized, not all magazines were available for the complete year 2019. When issues were not available for 2019, magazines were supplemented with issues from the corresponding month from the year 2020. The majority of the magazines had a monthly publishing frequency. However, *Black EOE Journal* published triannually; *Runner’s World* and *Taste of Home* published bimonthly; *The New Yorker, Woman’s World, Vanidades Mexico* and *Sports’ Illustrated* published weekly; *Saber Vivir Ar* published biweekly. For the magazines with weekly, biweekly, and fortnightly issues, the first issue of the month was chosen.

### Data collection

For each issue, the following information was recorded: magazine title, magazine type, issue, page number the advertisement was found on, advertisement category, product name, size of the advertisement, description of the model(s) in the advertisement if applicable (including number of models, gender, age, and race of model(s)), presence of a celebrity, magazine frequency, and number of monthly subscriptions. The advertising categories were recorded as exercise, fitness/exercise clothing, fitness/exercise equipment, health food, pain relief, supplements, diet, health insurance, weight loss programs, sports drink, medical establishment (diagnostic center), medical procedure, and medical equipment. If a certain advertisement product overlapped between two of the above advertisement categories, then the category that was primarily featured in the advertisement was chosen.

The 24 magazines were each reviewed independently by two individuals (AW, RO). Consensus was obtained by the examiners. If consensus was not reached, a third examiner (DW) was utilized to reach a final determination.

### US demographics

US racial demographics were used to control for racial variation in candidate analyses. Racial breakdowns were as follows: non-Hispanic White: 60.1%; non-Hispanic African American: 13.4%; Hispanic: 18.5%; Asian: 5.9%; Mixed race: 2.8% [24].

### Statistical analyses

Categorical variables were analyzed using Pearson’s chi-squared tests. When >20% of cells had expected frequencies <5, Fisher’s exact test was used. For intra-ad categories and intra-magazine type analyses, unknown race was dropped, and chi-squared Goodness of Fit tests were used. Goodness of fit analyses used distribution of current US racial demographics (see above). Only analyses with n≥10 were reported on and considered. Statistical analyses were performed using Stata v.16.1 and graphs were created using GraphPad Prism 7.0. Statistical significance was defined as p<0.05.

## Results

### MSK advertisements display demographic differences

A total of 1,617 MSK advertisements were identified. 832 (51.5%) advertisements did not display an identifiable model. Of the 785 ads with models, 538 (68.5%) featured a white model, 138 (17.6%) featured an African American model, 22 (2.8%) featured a Hispanic model, 10 (1.3%) featured an Asian model, 53 (6.8%) featured a model of mixed race, and 24 (3.1%) featured a model of unknown race or ethnicity (Table 1). This deviated significantly from US demographics (p<0.001). 57 (7.3%) featured multiple models of different races or ethnicities, while 728 (92.7%) did not (Table 1). Overall, white models were featured significantly more frequently than African American, Hispanic, and mixed race/ethnicity models (all p<0.001).

**Table 1.** Magazine and ad characteristics.

| Variable | Number MSK ads across 3 magazines/type; N (%) |
| --- | --- |
| **Magazine type** |  |
| Senior Health | 131 (8.11%) |
| Women's Health | 352 (21.78%) |
| Sports Health | 401 (24.81%) |
| Men's Health | 355 (21.97%) |
| Food/Dietary Health | 85 (5.26%) |
| General Lifestyle | 211 (13.06%) |
| African American Health | 12 (0.74%) |
| Hispanic Health | 69 (4.27%) |
| **Ad category** |  |
| Exercise | 173 (10.71%) |
| Fitness/Exercise Clothing | 250 (15.47%) |
| Fitness/Exercise Equipment | 237 (14.67%) |
| Health Food | 264 (16.34%) |
| Pain Relief | 150 (9.28%) |
| Supplements | 225 (13.92%) |
| Diet | 199 (12.31%) |
| Health Insurance | 19 (1.18%) |
| Weight Loss Program | 13 (0.80%) |
| Sports Drink | 62 (3.84%) |
| Medical Establishment | 6 (0.37%) |
| Medical Procedure | 2 (0.12%) |
| Medical Equipment | 16 (0.99%) |
| **Featured model** |  |
| Model | 785 (48.55%) |
| No model | 832 (51.45%) |
| **Multiple races in ad** |  |
| Yes | 57 (3.53%) |
| No | 1,560 (96.47%) |
| **Different genders in ad** |  |
| Yes | 133 (8.23%) |
| No | 1,483 (91.71%) |
| Unknown | 1 (0.06%) |
| **Race of model** |  |
| Black | 138 (17.58%) |
| Hispanic | 22 (2.80%) |
| Asian | 10 (1.27%) |
| Mixed | 53 (6.75%) |
| White | 538 (68.54%) |
| Unknown or N/A | 24 (3.06%) |
| **Featured gender** |  |
| Female | 434 (55.29%) |
| Male | 287 (36.56%) |
| Unknown or N/A | 1 (0.13%) |
| Both | 63 (8.03%) |
| **Featured gender and race** |  |
| White Men | 196 (11.88%) |
| White Women | 313 (18.97%) |
| Black Men | 50 (3.03%) |
| Black Women | 64 (3.88%) |
| Hispanic Men | 12 (0.73%) |
| Hispanic Women | 6 (0.36%) |
| Asian Men | 4 (0.24%) |
| Asian Women | 5 (0.30%) |
| Mixed Race Men | 15 (0.91%) |
| Mixed Race Women | 36 (2.18%) |
| Unknown or none | 949 (57.52%) |

**Table 2.** Differences in race of model by MSK ad category.

| Variable | N (%) |  |  |  |  |  |  |
| --- | --- | --- | --- | --- | --- | --- | --- |
|  | White | Hispanic | Asian | Mixed | Black | Unknown | p-value |
| **Ad category** |  |  |  |  |  |  | **<0.001** |
| Exercise | 93 (67.9%) | 10 (7.3%) | 2 (1.5%) | 11 (8.0%) | 17 (12.4%) | 4 (2.9%) |  |
| Fitness/Exercise Clothing | 97 (72.9%) | 2 (2.3%) | 3 (2.3%) | 7 (5.3%) | 21 (15.8%) | 3 (2.3%) |  |
| Fitness/Exercise Equipment | 67 (75.3%) | 2 (2.3%) | 3 (3.4%) | 8 (9.0%) | 9 (10.1%) | 0 (0.0%) |  |
| Health Food | 49 (69.0%) | 0 (0.0%) | 0 (0.0%) | 1 (1.4%) | 17 (23.9%) | 4 (5.6%) |  |
| Pain Relief | 53 (85.5%) | 0 (0.0%) | 1 (1.6%) | 3 (4.8%) | 4 (6.5%) | 1 (1.6%) |  |
| Supplement | 96 (81.4%) | 2 (1.7%) | 1 (0.9%) | 11 (9.3%) | 7 (5.9%) | 1 (0.9%) |  |
| Diet | 61 (78.2%) | 1 (1.3%) | 0 (0.0%) | 8 (10.3%) | 8 (10.3%) | 0 (0.0%) |  |
| Health Insurance | 7 (41.2%) | 0 (0.0%) | 0 (0.0%) | 0 (0.0%) | 10 (58.8%) | 0 (0.0%) |  |
| Weight Loss Program | 2 (25.0%) | 0 (0.0%) | 0 (0.0%) | 0 (0.0%) | 6 (75.0%) | 0 (0.0%) |  |
| Sports Drink | 0 (0.0%) | 0 (0.0%) | 0 (0.0%) | 4 (7.4%) | 39 (72.2%) | 11 (20.4%) |  |
| Medical Establishment | 0 (0.0%) | 5 (100.0%) | 0 (0.0%) | 0 (0.0%) | 0 (0.0%) | 0 (0.0%) |  |
| Procedure | 2 (100.0%) | 0 (0.0%) | 0 (0.0%) | 0 (0.0%) | 0 (0.0%) | 0 (0.0%) |  |
| Medical Equipment | 10 (100.0%) | 0 (0.0%) | 0 (0.0%) | 0 (0.0%) | 0 (0.0%) | 0 (0.0%) |  |

Bold indicates statistical significance, set at p<0.05.

**Table 3:** Observed frequencies of racial distribution within MSK ad categories differ from US demographic distribution.

| Variable | Observed frequency; expected frequency |  |  |  |  |  |
| --- | --- | --- | --- | --- | --- | --- |
|  | White | Hispanic | Asian | Mixed | Black | p-value |
| **Ad category** |  |  |  |  |  |  |
| Exercise | 93; 79.9 | 10; 24.6 | 2; 7.9 | 11; 3.7 | 17; 17.8 | **<0.001** |
| Fitness/Exercise Clothing | 97; 78.1 | 2; 24.1 | 3; 7.7 | 7; 3.6 | 21; 17.4 | **<0.001** |
| Fitness/Exercise Equipment | 67; 53.5 | 2; 16.5 | 3; 5.3 | 8; 2.5 | 9; 11.9 | **<0.001** |
| Health Food | 49; 40.3 | 0; 12.4 | 0; 4.0 | 1; 1.9 | 17; 9.0 | **<0.001** |
| Pain Relief | 53; 36.7 | 0; 11.3 | 1; 3.6 | 3; 1.8 | 4; 8.2 | **<0.001** |
| Supplement | 96; 70.3 | 2; 21.7 | 1; 6.9 | 11; 3.3 | 7; 15.7 | **<0.001** |
| Diet | 61; 46.9 | 1; 14.4 | 0; 4.6 | 8; 2.2 | 8; 10.5 | **<0.001** |
| Health Insurance | 7; 10.2 | 0; 3.2 | 0; 1.0 | 0; 0.5 | 10; 2.3 | **<0.001** |
| Weight Loss Program* | 2; 4.8 | 0; 1.5 | 0; 0.5 | 0; 0.2 | 6; 1.1 | **<0.001** |
| Sports Drink | 0; 25.8 | 0; 8.0 | 0; 2.5 | 4; 1.2 | 39; 5.8 | **<0.001** |
| Medical Establishment* | 0; 3.0 | 5; 0.9 | 0; 0.3 | 0; 0.1 | 0; 0.7 | **<0.001** |
| Procedure* | 2; 1.2 | 0; 0.4 | 0; 0.1 | 0; 0.1 | 0; 0.3 | 0.728 |
| Medical Equipment | 10; 6.0 | 0; 1.9 | 0; 0.6 | 0; 0.3 | 0; 1.3 | **0.037** |

\*n<10 so no significant statistical conclusions could be drawn. Bold indicates statistical significance, set at p<0.05.

### Racial demographics of model by MSK advertisement category

Significant differences in race/ethnicity were observed by MSK ad category (p<0.001) (Table 5). In all categories, white models were significantly overrepresented, apart from sports drink and health insurance advertisements, where they were underrepresented (all p<0.001, medical equipment p=0.037) (Table 6). African American models were overrepresented in health food, sports drinks, and fitness/exercise clothing, and underrepresented across pain relief and supplements (Table 6). Hispanic models were underrepresented across all ad categories (Table 6). Asian models had little to no representation, and were underrepresented across all categories (Table 6). Mixed race models were overrepresented in exercise, exercise and fitness clothing, equipment, pain relief, supplement, diet, and sport drink advertisements (Table 6).

## Discussion

In the U.S., significant racial and ethnic disparities exist in MSK health, as evidenced by differences in rates of obesity, hip and knee osteoarthritis, severe joint pain, and lost QALYs [1–7, 25]. Direct-to-consumer advertising is highly influential in minority communities [21, 22], and race portrayal in advertising has been shown to affect customer perception, favorability, and buying patterns [13–19, 21, 22]. It has been demonstrated that African Americans and Hispanics are underrepresented in health-promoting magazine advertisements [11, 12, 20, 22, 26]. If this lack of diversity also exists in MSK magazine advertising, a shift in representation could be an important mechanism to reduce disparities in MSK health. To this end, we sought to examine racial and ethnic representation in MSK advertisements. We collected data from 24 magazines across 8 magazine types.

Our study is limited by the difficulty and subjective nature of identifying models by race/ethnicity. For example, differentiating between hair color, facial features and skin color can sometimes be ambiguous. However, the impact of this ambiguity was mitigated by utilizing 2 independent racially diverse reviewers and adjudicating disagreements by a third reviewer. Additionally, this subjectivity in identifying models by race/ethnicity is also present when a viewer is exposed to an advertisement.

Overall, we found unbalanced racial representation across all MSK advertisements. We found that white models were represented significantly more than all other races, and that this representation proportionately deviated from US demographics. Despite white populations having lower rates of obesity [5], food insecurity [8], osteoarthritis [1–5], and overall better general health, white individuals were the most targeted audience for MSK products, even when adjusting for US racial demographics. We also found that white models were overrepresented in all ad categories, apart from sports drink and health insurance advertisements, where they were underrepresented.

In terms of African American models, they were overrepresented in sports drink advertisements and in sports health magazines. This reinforces previous literature, demonstrating the skewed presentation of African Americans as athletes [11]. This narrow representation may serve to further reduce the target audience for MSK advertisements in the African American community, perpetuating stereotypes and disparities. In addition, we found African American models were underrepresented across pain relief advertisements. Racial disparities in pain management advertising are concerning, given healthcare’s historical and current underdiagnoses of pain and treatment biases in the African American community [28, 29].

In terms of a positive and encouraging findings, health food and health insurance were two ad categories in which African Americans were over-represented, and these may potentially have a positive impact in addressing African American healthcare disparities. The increased inclusion of African American models in health food advertisements may serve to address high levels of food insecurity in African American communities. In addition, the increased representation in health insurance advertisements is also positive, given the racial disparities that exist as it pertains to health insurance coverage. As African American and Hispanic individuals have lower rates of health insurance coverage than non-Hispanic White individuals [27], this overrepresentation in insurance advertisements can be seen to be appropriately targeting a population in need, and a strategy that can be emulated in other categories.

Potential avenues to increase diversity include encouraging advertisers who did not utilize models to include a racially diverse model through hiring. For example, more than half of MSK advertisements did not have any model featured or displayed. This represents a straightforward actionable opportunity in which current ads without models could be revised to include models. Another alternative is to increase diversity in hiring of models in companies that utilized a disproportionate number of white models. Additionally, magazine publishers could provide guidance in terms of which populations would best benefit from learning about certain advertisement categories, encouraging advertisers to feature more minority models and target minority audiences. This may be an important step towards improving the observed racial and ethnic disparities in MSK health.

## Data Availability

The data underlying the results presented in the study are available from the corresponding author, Kelsey Rankin.

## Acknowledgements

N/A

## References

1. Allen, K.D., Racial and ethnic disparities in osteoarthritis phenotypes. Curr Opin Rheumatol, 2010. 22(5): p. 528–32.

2. Dillon, C.F., et al., Prevalence of knee osteoarthritis in the United States: arthritis data from the Third National Health and Nutrition Examination Survey 1991-94. J Rheumatol, 2006. 33(11): p. 2271–9.

3. Nelson, A.E., et al., Characterization of individual radiographic features of hip osteoarthritis in African American and White women and men: the Johnston County Osteoarthritis Project. Arthritis Care Res (Hoboken), 2010. 62(2): p. 190–7.

4. Sowers, M., et al., Radiographically defined osteoarthritis of the hand and knee in young and middle-aged African American and Caucasian women. Osteoarthritis Cartilage, 2000. 8(2): p. 69–77.

5. Wright, N.C., et al., Self-reported osteoarthritis, ethnicity, body mass index, and other associated risk factors in postmenopausal women-results from the Women’s Health Initiative. J Am Geriatr Soc, 2008. 56(9): p. 1736–43.

6. Losina, E., et al., Impact of obesity and knee osteoarthritis on morbidity and mortality in older Americans. Ann Intern Med, 2011. 154(4): p. 217–26.

7. Vaccaro, J.A. and F.G. Huffman, Sex and Race/Ethnic Disparities in Food Security and Chronic Diseases in U.S. Older Adults. Gerontol Geriatr Med, 2017. 3: p. 2333721417718344.

8. Larson, N.I., M.T. Story, and M.C. Nelson, Neighborhood environments: disparities in access to healthy foods in the U.S. Am J Prev Med, 2009. 36(1): p. 74–81.

9. August, K.J. and D.H. Sorkin, Racial/ethnic disparities in exercise and dietary behaviors of middle-aged and older adults. J Gen Intern Med, 2011. 26(3): p. 245–50.

10. Bowen l and S. J, Minority Presence and Portrayal in Mainstream Magazine Advertising: An Update. Journalism & Mass Communication, 1997. 74(1): p. 134–146.

11. Plous, S. and D. Neptune, Racial and Gender Biases in Magazine Advertising: A ContentAnalytic Study. Psychology of Women Quarterly, 1997. 21(4).

12. Appiah, O. Black, White, Hispanic, and Asian American Adolescents’ Responses to Culturally Embedded Ads. Howard Journal of Communication, 2001. 12(1): p. 29–48.

13. Bush, R.F., R.F. Gwinner, and P.J. Solomon, White Consumer Sales Response to Black Models. Journal of Marketing, 1974. 38(2): p. 25–29.

14. Chaiken, S., Heuristic versus Systematic Information Processing and the Use of Source versus Message Cues in Persuasion. Journal of Personality and Social Psychology, 1980. 39(5): p. 752–766.

15. Cui, G., Marketing Strategies in a Multi-Ethnic Environement. Journal of Marketing Theory and Practice, 1997. 5(1): p. 122–134.

16. Davis, J.L. and O.H. Gandy, Racial Identity and Media Orientation: Exploring the Nature of Constraint. Journal of Black Studies, 1999. 29(3): p. 367–397.

17. Deshpande, R., W.D. Hoyer, and N. Donthu, The Intensity of Ethnic Affiliation: A Study of the Sociology of Hispanic Consumption. Journal of Consumer Research, 1986. 13(2): p. 214–220.

18. Grier, S.A., A.M. Brumbaugh, and C.G. Thornton, Crossover Dreams: Consumer Responses to Ethnic-Oriented Products. Journal of Marketing, 2006. 70(2): p. 35–51.

19. Duerksen, S.C., et al., Health disparities and advertising content of women’s magazines: a cross-sectional study. BMC Public Health, 2005. 5: p. 85.

20. Media, A.o.A., Circulation Report. 2019.

21. Bureau, U.S.C., Quick Facts: United States. 2021.

22. Kulkarni, K., et al., Obesity and osteoarthritis. Maturitas, 2016. 89: p. 22–8.

23. Prevention, C.f.D.C.a., Racial/ethnic differences in the prevalence and impact of doctor diagnosed arthritis. MMWR, 2005. 54(5): p. 119–123.

24. Aaker, J.L., A.M. Brumbaugh, and S.A. Grier, Nontarget Markets and Viewer Distinctiveness: The Impact of Target Marketing on Advertising Attitudes. Journal of Consumer Psychology, 2000. 9(3): p. 127–140.

25. Lee, D. and C.E. Begley, Racial and ethnic disparities in response to direct-to-consumer advertising. Am J Health Syst Pharm, 2010. 67(14): p. 1185–90.

26. Wiznia, L.E., et al., Deficiency of sun protection advertising exists in consumer magazines across demographic groups and varies by target demographic. J Am Acad Dermatol, 2019. 80(4): p. 1139–1141.

27. Hoffman, K.M., et al., Racial bias in pain assessment and treatment recommendations, and false beliefs about biological differences between blacks and whites. Proc Natl Acad Sci U S A, 2016. 113(16): p. 4296–301.

28. Maina, I.W., et al., A decade of studying implicit racial/ethnic bias in healthcare providers using the implicit association test. Soc Sci Med, 2018. 199: p. 219–229.

29. Artiga, S., K. Orgera, and A. Damico, Changes in Health Coverage by Race and Ethnicity since the ACA, 2010-2018,. Kaiser Family Foundation, 2020.

